# Descriptive epidemiology of COVID-19 deaths during the first wave of pandemic in India - a single center experience

**DOI:** 10.1101/2021.09.01.21262791

**Authors:** Prakash Tendulkar, Pragya, Prasan Kumar Panda, Ajeet Singh Bhadoria, Poorvi Kulshreshtha

## Abstract

**Background:** With the looming threat of recurrent waves of COVID-19 in the presence of mutated strains, it’s of paramount importance to understand the demographic and clinical attributes of COVID-19 related mortalities in each pandemic waves. This could help policy makers, public health experts, and clinicians to better plan preventive and management strategies to curb COVID-19 related mortality.

**Method:** This was a hospital record based, retrospective cross-sectional descriptive study, at a tertiary care hospital in Rishikesh, India. The study included all deceased patients between March 2020 and January 2021 (first wave) who tested positive for SARS-CoV-2 by RT-PCR and were hospitalized. The study was done to describe demography, clinical presentation, laboratory parameters, treatment given and associated complications of all COVID-19 deaths.

**Result:** Out of 424 mortalities, 298 (70.38%) were males and 126 (29.62%) were females. Mean age of patients was 55.85 ± 16.24 years, out of which 19.5 percent were less than 45 years old, 33.6 percent were 45 to 60 years old and 41.8% were more than 60 years old. Comorbidity in the form of type 2 diabetes mellitus was present in 41.4% [95% CI (41.4-51.1)], hypertension in 39.8% [95% CI (35.1-44.6)], and coronary artery disease in 15.2% [95% CI (11.8-18.8)]. At the time of presentation, shortness of breath was present in 73.6% [95% CI (69.1 -77.7)], fever in 64.92% [95% CI (60.1-69.4)], and cough in 46.1%, [95% CI (41.1-50.8)]. Deranged laboratory parameters were lymphopenia in 90.2% [95% CI (86.8-92.7)], transaminitis in 59.7% [95% CI (54.8-64.3)], and hypercreatinemia in 37.7% [95% CI (33.1-42.5)]. Complications manifested were acute respiratory distress syndrome in 78.3% [95% CI (74-82.1)] and shock in 54.7% [95% CI (49.8-59.5)]. Median time duration between onset of symptom and hospital admission was 5 days (IQR = 3 - 5 days) and median length of hospital stay was 9 days (IQR = 4 - 14 days).

**Conclusion:** During first pandemic wave, COVID-19 related mortality was 2.37 times higher among males, 2.14 times in age group >60 than <45 years. Most common associated comorbidities (>40%) were type 2 diabetes mellitus and hypertension. Most common associated symptoms (>60%) were shortness of breath and fever. Lymphopenia was seen in >90% cases while liver involvement in 60% and kidney in 38% cases. Median hospital stay was doubled the pre-hospital illness.

## Introduction

COVID-19 was declared a global pandemic by the World health organization (WHO) on Mar 11 2020. As of August 15 2021, a total of 207,542,620 confirmed cases were diagnosed all over the world, with a total death of 4,367,396 (WHO data on COVID-19). India has reported over 32 million coronavirus cases as of August 14, 2021 with more than 31 million recoveries and around 430 thousand causalities while Uttarakhand, an Indian state in Himalayan region, reports over 340 thousand confirmed cases and 7370 deaths (MOHFW, India data on COVID-19).

From January 2020 till February 2021 of first wave effects, September 2020 reported highest number of cases and causalities in India (MOHFW, India data on COVID-19). Seeing the dynamic nature of coronavirus, it’s of utmost importance to understand the demographic and clinical attributes of COVID related mortalities during each waves. This could help policy makers and clinicians to plan ahead to curb COVID-19 deaths.

This is a single centre cross-sectional study to describe the clinical characteristic of deceased COVID-19 patients admitted at a tertiary care hospital, Rishikesh.

## Method

This was a hospital record based, retrospective descriptive study. The study includes all deceased patients who tested positive for SARS-CoV-2 by RT-PCR between March 2020 and January 2021 and were hospitalized for COVID-19 illness. Data was obtained from patient records available at the e-hospital, Health Information Management System portal of the Government of India. The study was done to assess risk factors, comorbidities, clinical signs and symptoms, laboratory parameters, treatment course, associated complications, the time interval between symptom onset and hospital admission as well as time interval between hospital admission and death.

Appropriate ethical approval was obtained from the institute’s ethical committee, AIIMS, Rishikesh before assessing the required data. Data management was done on Microsoft excel spreadsheet. Categorical variables were described as frequency and proportion. Continuous variables described as mean ± standard deviation or median with inter-quartile range (IQR) as applicable.

## Results

Out of the total reported COVID-19 cases in Uttarakhand till January 2021 (first wave), 2396 got admitted at this tertiary care hospital and 424 causalities reported. Out of this, 297 (70.3%) were males, and 125 (29.7%) were females. The mean age in years was 55.85 ± 16.24 years. out of which 19.5 percent were less than 45 years old, 33.6 percent were 45 to 60 years old and 41.8% were more than 60 years old.

The most common symptoms among the deceased were shortness of breath [73.6%, 95% CI (69.1-77.7)], fever [64.92%, 95% CI (60.1-69.4)], and cough [46.1%, 95% CI (41.1-50.8)]. Other less common symptoms include sore throat [12.5%, 95% CI (9.98-16.43)] chest pain [11.2%, 95% CI (8.51-14.59)], altered mental status [9.8%, 95% CI (7.4-13.28)], abdominal pain [6.5%, 95% CI [4.2-8.9)], diarrhoea [6.3%, 95% CI (4.04-8.69)], headache [2.2%, 95% CI (0.09-3.80)], loss of taste and smell [1.1%, 95% CI (0.15-2.20)] (Figure1).

**Figure 1:**
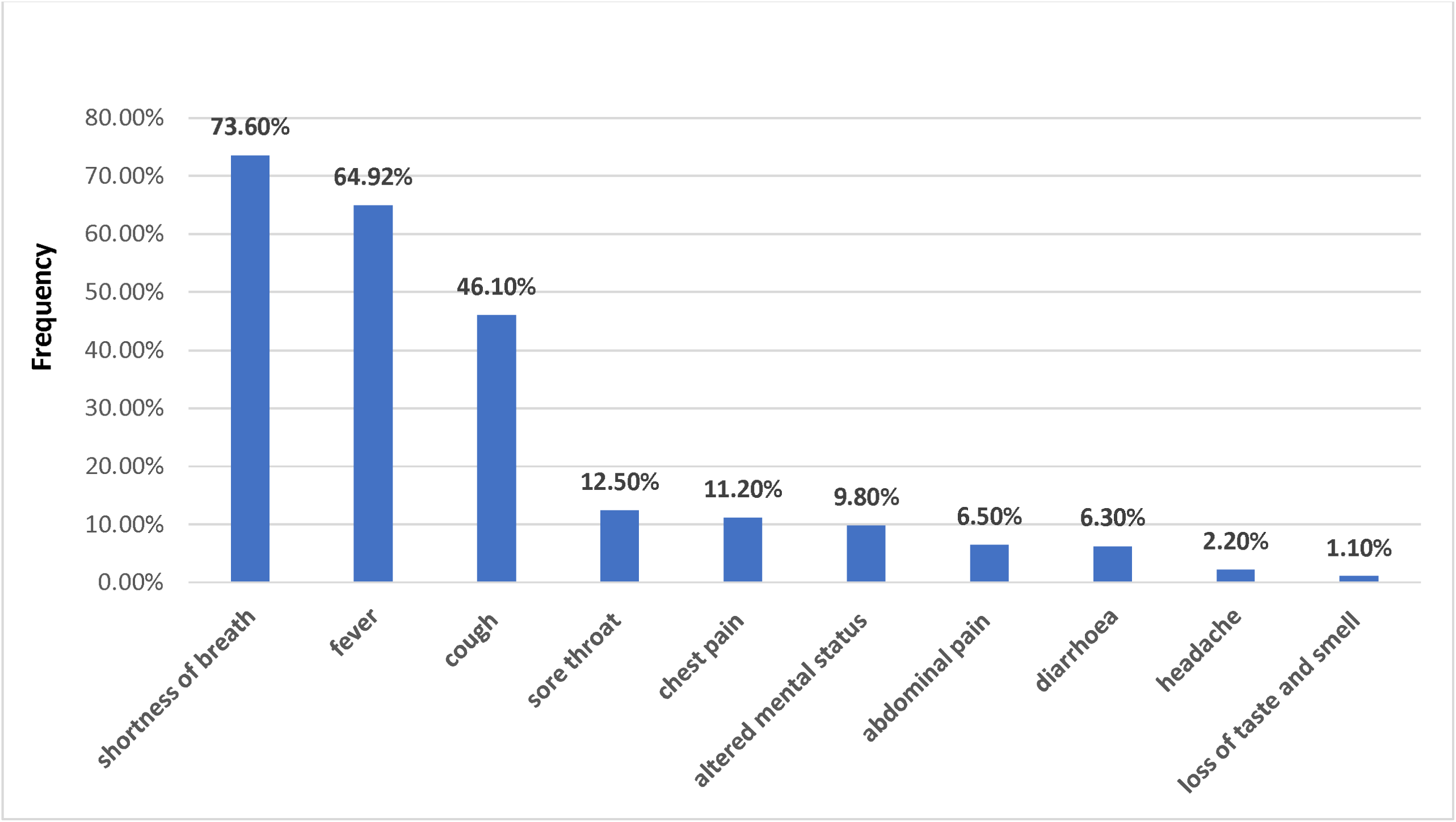
Symptoms in deceased COVID-19 patients.

Tachypnoea (RR>20) was present in 339 [80%, 95% CI (76.14-83.76)] of the patient. 275, [64.86 %, CI (60.10-69.40)] patients had hypoxia and needed oxygen support at admission i.e., severe COVID-19. 36 [8.1%, 95% CI (5.83-11.14)] of the patient had hypotension at admission.

In comorbidity, diabetes [41.4%, 95% CI (41.4-51.1)] was the most common. Other comorbidities were hypertension [39.8%, 95% CI (35.1-44.6)], coronary artery disease (CAD) [15.2%, 95% CI (11.8-18.8)], chronic obstructive airway disease (COPD) [8.3%, 95% CI (6.04-11.41)], chronic kidney disease (CKD) [7.6%, 95% CI (5.43-10.60)], malignancy [5.4%, 95% CI (3.26-7.58)], chronic neurological disorder was present in 3.6% [95% CI (1.77-5.29)] and chronic liver disease (CLD) [(3.4%, 95% CI (1.60-5.00)] (Figure 2). 188 patients (44%, 95% CI (39.55-49.21%)] had ≥ 2 of these co morbidities. Out of all deceased 7 [1.65 %, 95% CI (0.67 – 3.37 %)] were pregnant.

**Figure 2:**
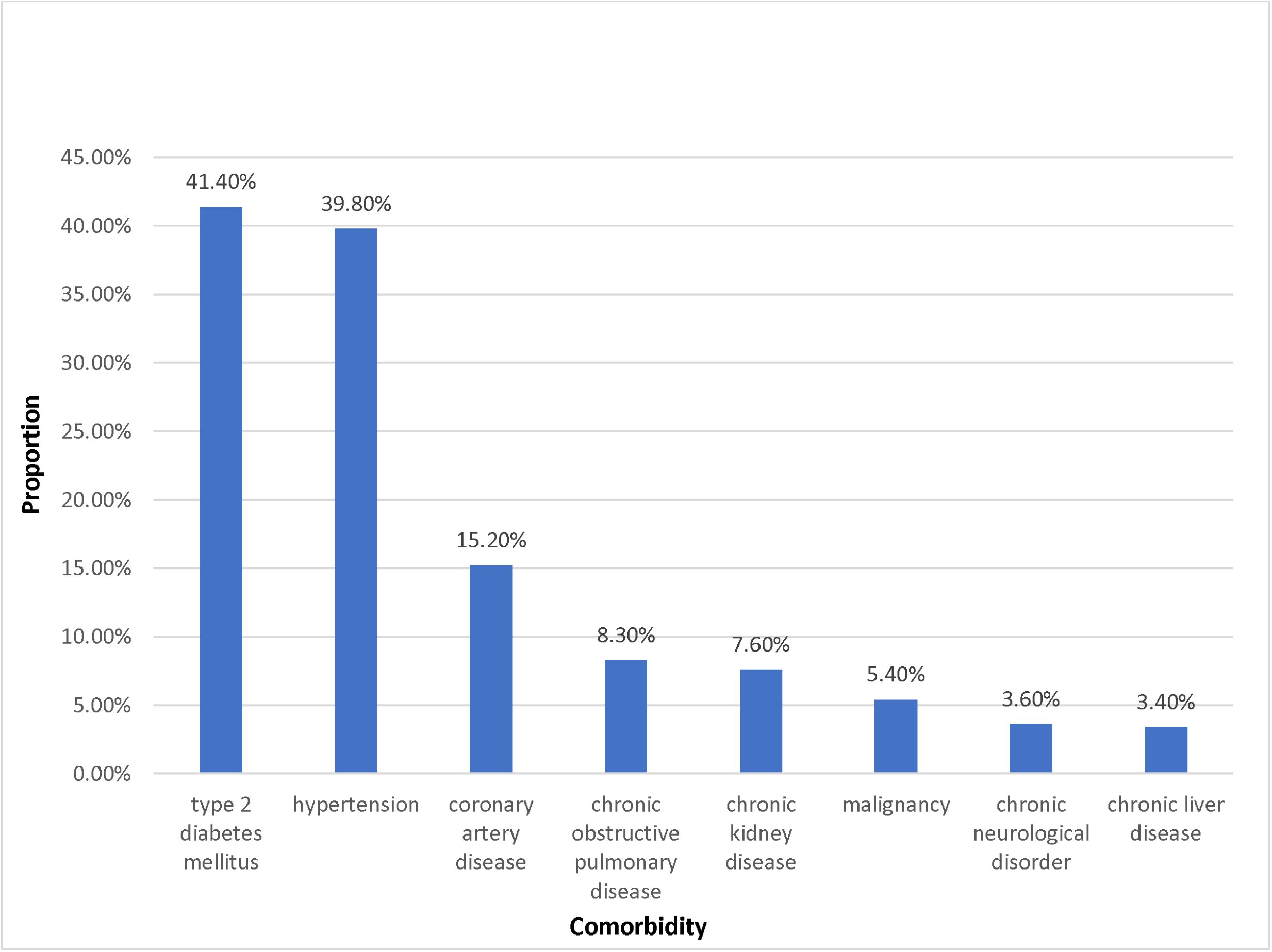
Comorbidity in deceased COVID-19 patients.

If we see in laboratory parameters at admission, lymphopenia was present in 403 [90.2%, 95% CI (86.8-92.7)] patients, leucocytosis in 214 [50.5%, 95% CI (45.71-55.23)], anaemia in 122 [28.8%, 95% CI (24.46-33.08)], thrombocytopenia in 112 [25.1%, 95% CI (22.21-30.61)] patients. 253 (59.7%, 95% CI (55.00-64.33)] patients had raised Alanine Aminotransferase (ALT) levels and raised Aspartate Aminotransferase (AST) level in 149 [35.1%, 95% CI (54.8-64.3)]. 160 [37.7%, 95% CI (33.1-42.5)] patient had creatinine of more than 1.2mg/dl (Figure 3).

**Figure 3:**
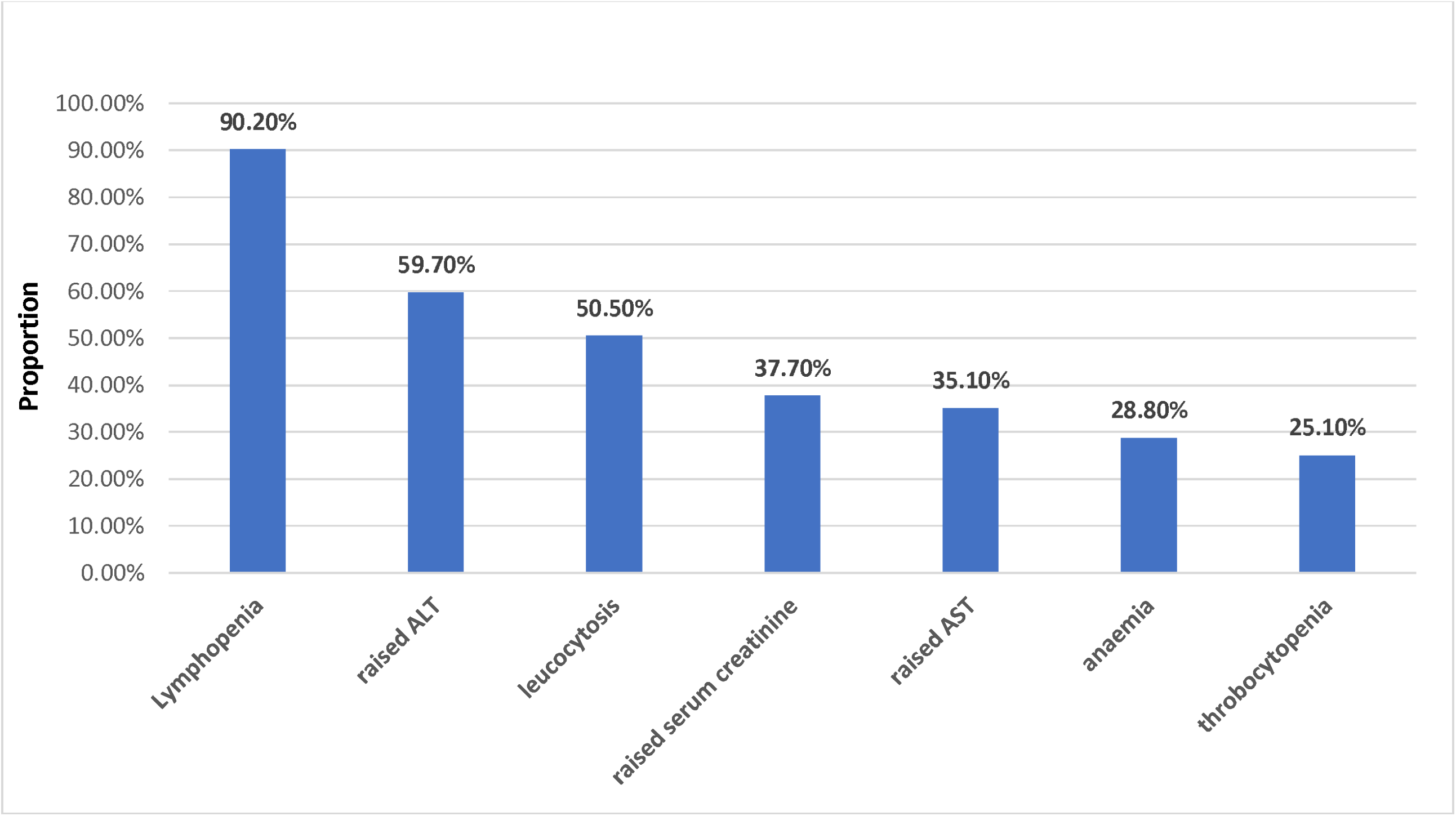
Lab parameters in deceased COVID-19 patients.

Three hundred thirty-two patients (78.3%, 95% CI) needed ICU facility at admission. 237 (55.9%, 95% CI) patients started on NIV support at admission. 232 [54.7%, 95% CI (49.8-59.5)] developed shock during the hospital stay and needed inotropic support. 46 (10.3%) patients needed renal replacement therapy during the hospital stay. 144 [33.96%, CI (29.5-38.5)] patients were managed with IV steroids. Acute Respiratory Distress Syndrome (ARDS) was present in 292 [68.87%, CI (64.22-73.25)] patients, sepsis was present in 171 patients [40.33%, CI (35.7-45.0)].

Median time duration between onset of symptom and hospital admission was 5 days (IQR = 3-8 days) and median length of hospital stay was 9 days (IQR = 4-14 days). (Figure 4).

**Figure 4A:**
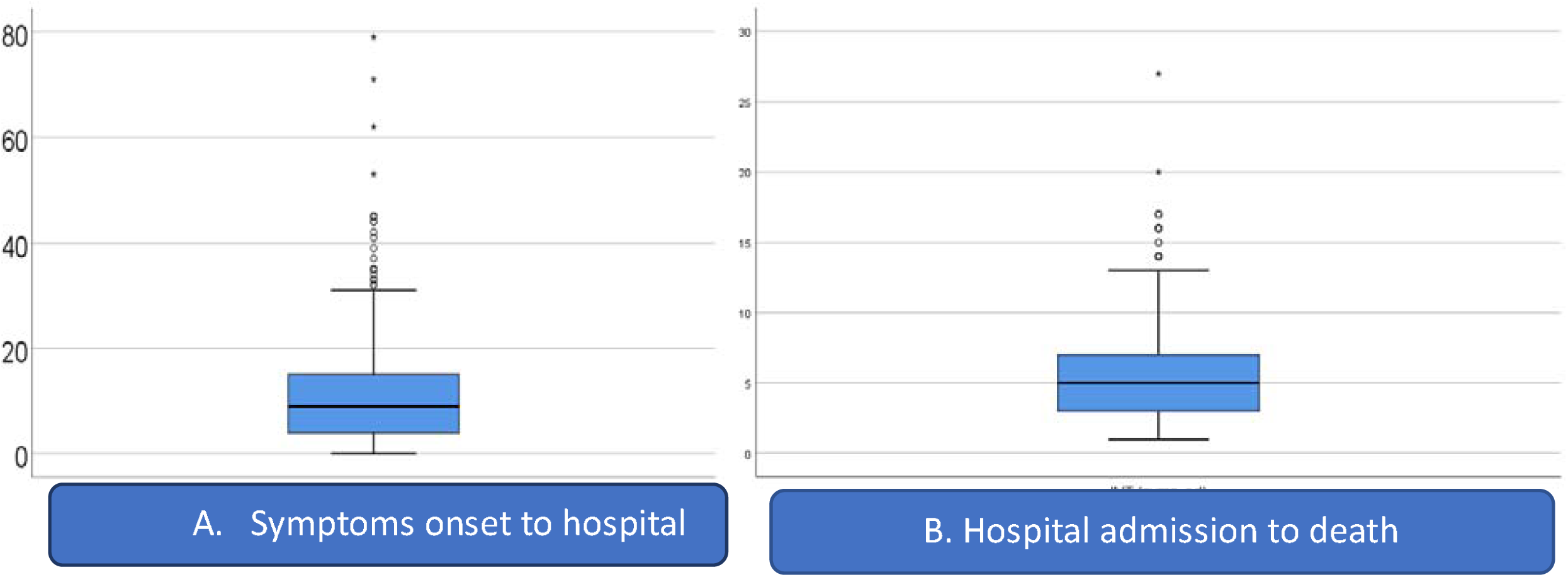
Duration (in days) from symptom onset to hospital admission, 4B: length of hospital stay (in days) in deceased COVID-19 patients.

## Discussion

As we are witnessing the current trend globally, mortality in the COVID-19 pandemic is very high. Strikingly, most deaths are seen in the male population than female.^(1)(2)^ Results of a hospital based COVID related death study from United States shows that 60.6% of the deceased were males.^(1)^ One review article supported the evidence that “ineffective anti-SARS-CoV-2 responses, coupled with a predisposition for inappropriate hyperinflammatory responses, could provide a biological explanation for the male bias in COVID-19 mortality”.^(3)^ The present study showed a similar trend, and out of total patients, 70.3% patients were males. Another explanation for higher mortality in males could be limited health care access to females leading to low footfalls in hospitals.^(4)^ In the present study, around 42% of deceased were more than sixty year old. Earlier studies have also suggested that mortality is higher in elderly males.^(1) (2) (5)(6)^

Comorbidity also plays an essential role in any disease related mortality. A study done on Italian deceased patients has shown that, 35.5% of the deceased had diabetes, 30% had ischemic heart disease, 20.3% had active cancer, 24.5% had atrial fibrillation, 6.8% had dementia, and 34 (9.6%) had a history of stroke.^(2)^ Results of another hospital-based study from southern India shows that diabetes (62%), hypertension (49.2%), and CAD (17.5%) were the commonly reported comorbidities.^(7)^ Our study also shows similar findings with regard to comorbidities with diabetes being the most common comorbidity followed by hypertension, CAD, COPD, CKD, malignancy, chronic neurological disease, and CLD. In our study, 188 (44%) of the deceased had two or more comorbidities.

Dyspnoea was the most common presenting complain in the present study which is consistent with the finding of other studies.^(8)(9)^ 73.6% had dyspnoea at presentation, and 78.3 % needed ICU admissions.

One of the studies in China showed that 83.2% of the patients had lymphopenia on admission.^(10)^ Another Korean nationwide study concluded that lymphopenia at the time of initial presentation is associated with poorer prognosis in COVID-19 patients.^(11)^ In the present study, 90.2% deceased patients had lymphopenia at the time of admission. One of the metanalyses showed that compared to moderate cases, severe COVID-19 cases had anaemia at presentation [weighted mean difference (WMD), - 4.08 g/L (95% CI - 5.12; - 3.05).^(12)^ 28.8% of the patients had a haemoglobin of ≤ 11 gm/dl at admission, The meta-analysis suggested that thrombocytopenia enhanced the risk of severe COVID-19 death by over fivefold.^(13)^

One previous retrospective study suggested that both pre-renal and intrinsic AKI were associated with higher mortality.^(14)^ Meta-analysis suggested that the risk of death in patients with AKI in COVID-19 increases significantly (OR 11.05, 95% CI (9.13-13.36)).^(15)^ One of the article demonstrated, out of 3993 patients COVID-19 positive patients, 1835 (46%) developed AKI and total of 347 (19%) required dialysis.^(16)^ In the present study, 37.7 % of patients had raised creatinine levels at admission. But, only 7.6% of patients were known for CKD, and the rest, 30.1% had AKI due to COVID-19.

The shock was also a complication in ICU patients. One meta-analysis suggested that in critically ill patients, 32% patients developed shock.^(17)^ In the present study study, 8.1% had a shock at admission, and 54.7% of patients needed inotropic support in the hospital.

ARDS is the leading cause of COVID related mortality. Previous systematic review and meta-analysis also supported this and ARDS was the most common complication, in both mild and severe patients.^(18)^ In the present study, 68.87 % of the deceased patients had ARDS complicating the disease.

The median duration of hospital stays in a study in China was 10-13 days, similar to the present study.^(10)(19)^ Similarly in one of the hospital based COVID related death study from United States, median interval from hospital admission to death was 5 days (IQR: 3, 8).^(1)^ Results of another hospital-based study from southern India shows the median time interval between symptom onset and hospital admission was 4 days (IQR: 2, 7); admission and death was 4 days (IQR: 2, 7) compared to 9 days in the present study.^(20)^

## Conclusion

In the Himalayan region of Uttarakhand, during the first wave of pandemic, a large proportion of RT-PCR positive COVID-19 deceased were elderly male with pre-existing comorbidities. Most common associated comorbidities were type 2 diabetes mellitus, hypertension and cardiovascular disease. Most common presenting complains were shortness of breath, fever, and cough. Most common deranged laboratory parameters are decreased lymphocyte count, elevated SGPT/ALT, and raised serum creatinine. Median pre-hospital illness was half the duration of hospital stay.

## Data Availability

After obtaining approval from corresponding author, de-identified data can be shared.

## Authors’ contributions

All authors made substantial contributions to conception and design, acquisition of data, or analysis and interpretation of data; took part in drafting the article or revising it critically for important intellectual content; agreed to submit to the current journal; gave final approval of the version to be published; and agree to be accountable for all aspects of the work.

## Ethics and Data sharing

The study was done after institute ethical approval and as per declaration of Helsinki. After obtaining approval from corresponding author, de-identified data can be shared.

## Acknowledgment

A gratitude to the entire COVID-19 management team during this pandemic crisis for helping in data collection.

